# Normal Blood Pressure in Healthy Adult Eritreans

**DOI:** 10.1101/2023.06.22.23291695

**Authors:** Ahmed O Noury, Barakat M Bakhit, Montasir A Osman, Omer A. Musa

## Abstract

**Background and objectives:** Blood pressure levels may vary because of genetic, ethnic, and socioeconomic factors. To date, there have been no large national studies in Eritrea on blood pressure. Therefore, we sought to establish a representative blood pressure reference interval for Health adult Eritreans

**Methods:** The study included a sample of 942 healthy Eritrean individuals aged between 18 and 60 years, comprising 331 males and 611 females. The participants were selected from households located in the cities of Asmara, Keren, and Mandafara. Anthropometric measurements and clinical data were collected from each participant. Blood pressure was measured twice using a standardized procedure.

**Results:** 942 healthy adults were included, 331 (35.1 %) were males with an average age of 40, and 611 (64.9%) were females, with an average age of 41, with an age range of 18-60 years. Median blood pressure for males was 120/78 mmHg, while the Median blood pressure for female was 118/78 mmHg for female

**Conclusion:** The values of blood pressure were similar to international values of blood pressure and there was significantly higher blood pressure in males than in females.

## Introduction

Blood pressure is a critical vital sign that guides both short- and long-term clinical decisions. Because blood pressure is so important in directing care, it must be measured accurately and consistently. During a blood pressure measurement, two values are typically recorded. The first, systolic pressure, is the maximum arterial pressure during systole. The second, diastolic pressure is the minimum arterial pressure during diastole. (1)

The American Heart Association (AHA) recommends the following blood pressure reference val ues (2):Normal: Less than 120/80 mm Hg, Elevated: Systolic between 120-129 mm Hg and diastolic less than 80 mm Hg, Stage 1 hypertension: Systolic between 130-139 mm Hg or diastolic between 80-89 mm Hg, Stage 2 hypertension: Systolic at least 140 mm Hg or diastolic at least 90 mm Hg, Hypertensive crisis: Systolic over 180 mm Hg and/or diastolic over 120 mm Hg. There are different definitions of hypertension. An SBP of 130 mmHg or higher, or a DBP of 80 mmHg or higher, is considered to be hypertension, according to the American College of Cardiology and the American Heart Association (3). The European Society of Cardiology and the European Society of Hypertension describe hypertension as having a DBP of 90 or higher or an SBP of 140 mmHg or higher(4).

It is important to understand that normal blood pressure levels might differ between populations due to a variety of factors like race and ethnicity. Compared to Caucasians, people of African descent have greater blood pressure and a higher chance of developing hypertension(5).Globally, the mean blood pressure and prevalence of hypertension differ significantly (6). Geographical latitude, ambient temperature, and the number of daylight hours have also been related to blood pressure and hypertension prevalence (7–9).

Therefore, establishing population-specific reference values is essential for accurate assessment and management of blood pressure. In the context of Eritrea, a country situated in the Horn of Africa, there is a paucity of data on normal blood pressure ranges in the local population. Eritrea is characterized by a unique combination of geographic factors and ethnic diversity, all of which can potentially influence blood pressure levels. The significance of investigating and establishing normal blood pressure values specific to Eritreans lies in its potential to enhance the precision of clinical assessments, facilitate early detection of hypertension, and support effective management strategies tailored to the local population. This study aims to address the gap in knowledge by investigating the influence of geographic factors, gender, and ethnicity on blood pressure patterns in healthy adults in Eritrea. By doing so, we aim to support clinical decision-making, facilitate early detection of hypertension, and promote effective preventive measures aligned with the unique characteristics of the Eritrean population.

### Methodology

## Materials and Methods

### Study design

This is descriptive and cross-sectional

### Study site

The study was conducted on Eritreans from all over the country in the period from 2019 up to 2022. Samples used in this study were collected from the following sites in Eretria:

Central region (Asmara)
Anseba region (Keren)
Southern region (Mendefera)
Northern Red Sea region (Massawa) Volunteer recruitment:

Participants were selected for the study using a multistage sampling technique (stratified sampling). The selection process involved several stages. In the first stage, a zone was randomly chosen from multiple zones in the city. In the second stage, a sub-zone within the selected zone was randomly chosen. Moving on to the third stage, blocks within the selected sub-zone were randomly chosen, with four blocks being selected in total. Finally, households were systematically chosen from a randomly selected reference point within the blocks. To ensure a diverse representation, the recruitment of participants was stratified into four age groups: 18-29, 30-39, 40-49, and 50-60 years. Prior to measuring blood pressure, written informed consent was obtained from all participants. The research team utilized a questionnaire to collect various data, including anthropometric measurements, demographic information, medical status, medical history, physical activity levels, and sleeping hours. Blood pressure and body mass index (BMI) measurements were then taken for all participants

### Reference population

This cross-sectional study included 942 healthy adult Eritreans from various social, ethnic, and professional groups. Our reference sample included 331 men with an average age of 40 and 611 women with an average age of 41; the study lasted from November 2019 to January 2022.

### Ethical clearance

The Eritrean Ministry of Health’s ethics committee granted ethical approval for this study.

### Blood pressure measurement

Blood pressure measurements were done by the auscultation method using mercury sphygmomanometers and stethoscopes. The patient is asked to sit in a comfortable chair. The arm is supported at heart level. A cuff of a suitable size is wrapped around the upper arm, about 1 inch above the bend of the elbow. The stethoscope is placed over the brachial artery, located on the inner side of the arm, about halfway between the elbow and the shoulder. The cuff is inflated to a pressure of about 30 mmHg above the patient’s expected systolic blood pressure. The cuff is slowly deflated while listening for the Korotkoff sounds. The systolic blood pressure is the first sound heard when the cuff is deflated. The diastolic blood pressure is the last sound heard before the sounds disappear. Two readings are taken at least 5 minutes apart using the auscultation method with a mercury sphygmomanometer and stethoscope. If the difference between the two readings is 10 mmHg or more for either SBP or DBP, a third reading is obtained. The final reading is the average of the two closest readings for SBP and DBP, or the nearest two readings if a third reading was obtained.

Statistical method: Moreover, this study makes use of a number of statistical methods. Mann-Whitney analysis was used to compare SBP and DBP between males and females(10). The Kruskal-Wallis test was also applied to look at how the study variables varied by area(11). In addition, scatter plots, boxplots, density plots, and histograms are employed for data visualization(12).

## Results

This section contains a presentation of the study’s findings. In addition to data visualization, the results of the Mann-Whitney test for assessing gender differences in the study variables (SBP and DBP) and the Kruskal-Wallis test for investigating differences by area are shown. Also included are descriptive statistics for the reference values.

As seen by the histogram and density in figure 1, SBP was concentrated around 120 in all locations, but with positive skewness in every instance. As the density represents, there are two picks at 110 and 120 in Eritrea as a whole. Similar to this, the data for DBP is gathered at 80 with a noticeable right skewness. Also, the chart shows that Asmara’s distribution of the two variables is more representative of Eritrea as a whole.

**Figure 1:**
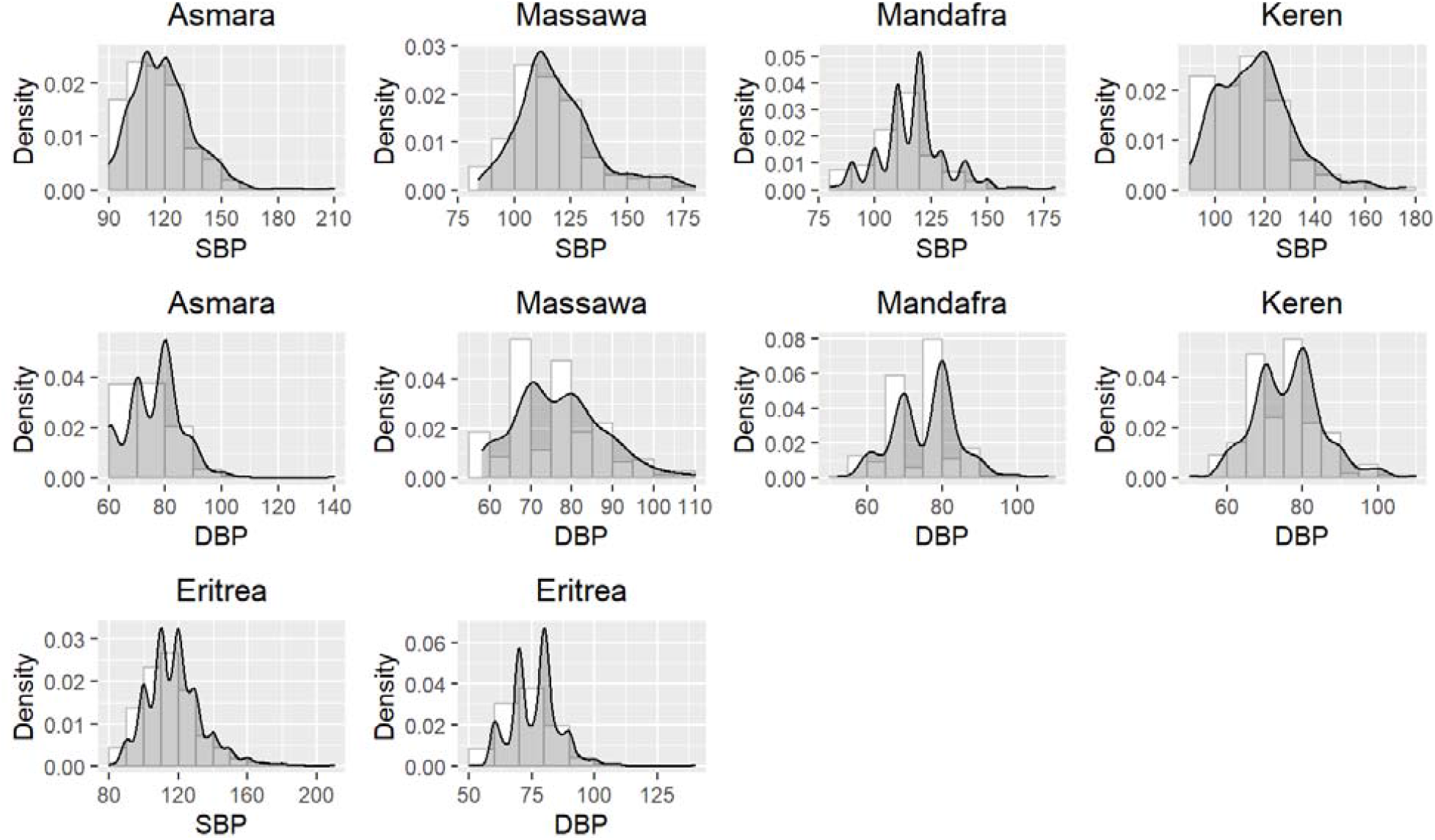
Histogram and density of SBP and DBP by region.

Figure 2’s boxplots of the study variables display the extreme values. The SBP has numerous higher extreme values in all locations, as shown in the figure, but the lower extreme values are smaller in Massawa and Keren and nil in Asmara, Mandafra, and Eritrea as a whole. Similar circumstances apply to DBP, where the highest extreme values are greater than the lower ones. DBP, on the other hand, has less extreme values than SBP.

**Figure 2:**
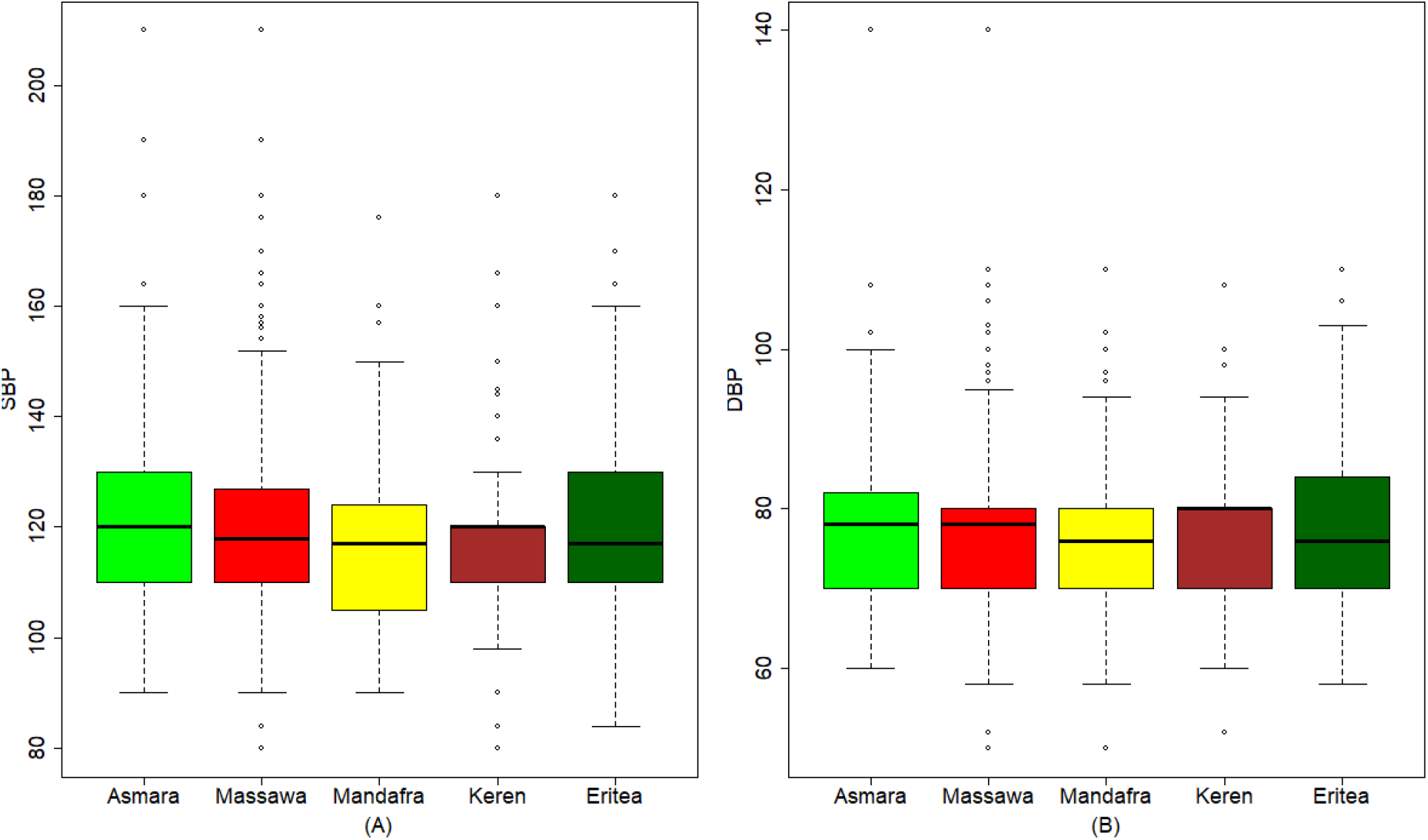
Boxplot of SBP and DBP by region.

In all regions, and especially in Massawa, the scatter plots in figure 3 demonstrate a positive linear correlation between SBP and age. In contrast, age has no bearing on DBP, with data distribution being constant from 20 to 60 years of age.

**Figure 3:**
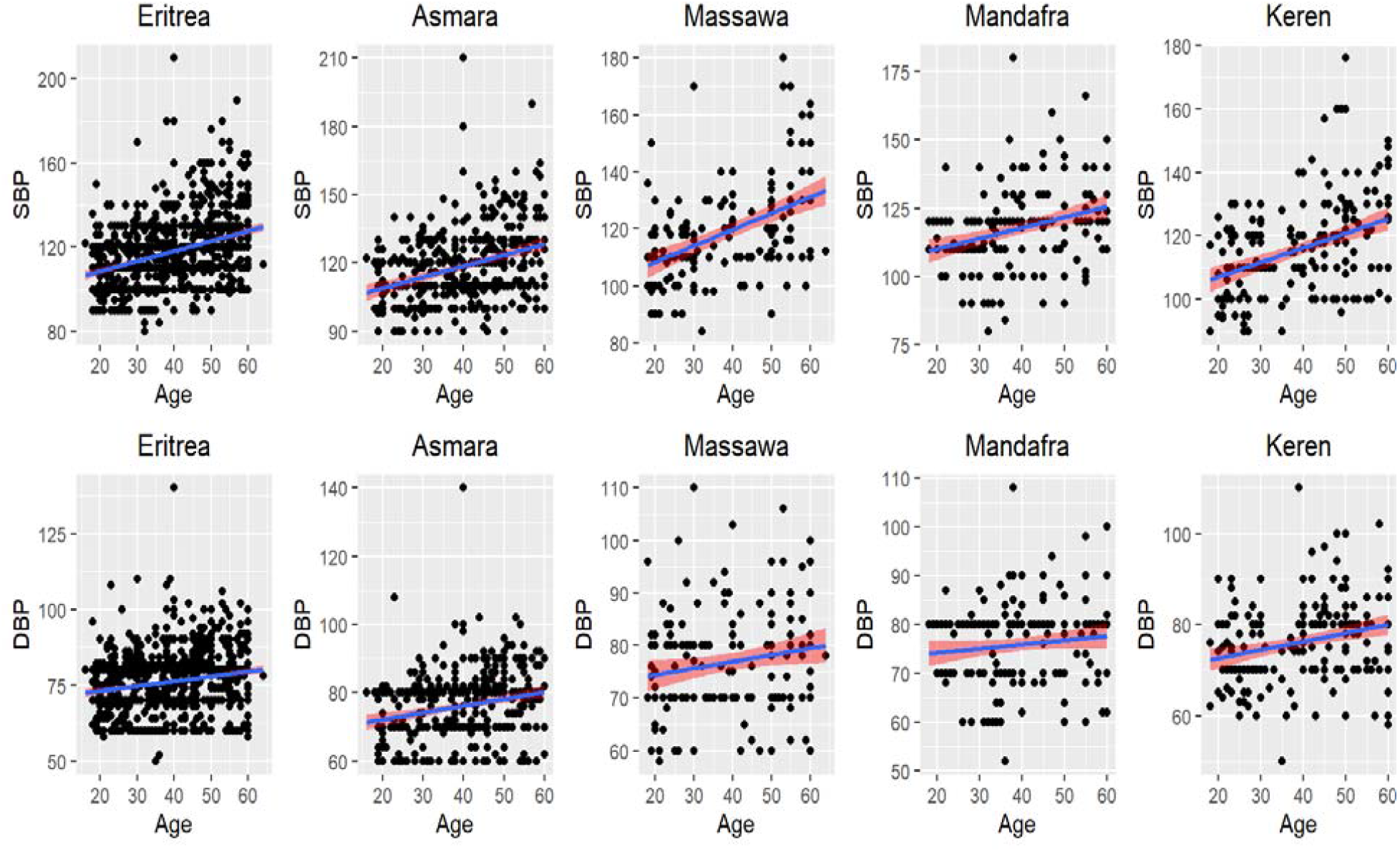
Scatter plot of the relationship between SBP and DBP and age by region.

**Figure 4:**
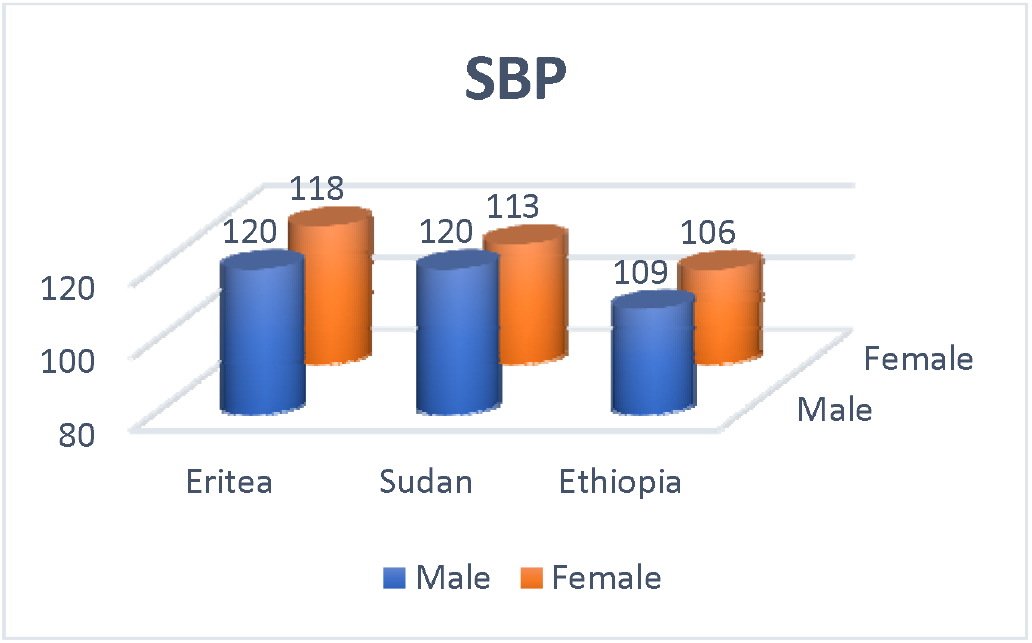
Comparison systolic blood pressure levels of Eritreans with values of Sudanese and Ethiopians

**Figure 5:**
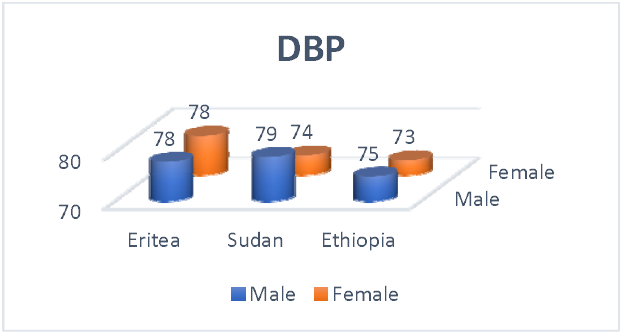
Comparison Diastolic blood pressure levels of Eritreans with values of Sudanese and Ethiopians

The results of the Mann-Whitney test to compare SBP and DBP by gender are shown in table 3. According to the findings, there are no statistically significant disparities between men and women in Asmara, Massawa, Mandafra, or Etritrea, however there is a difference in Keren (the p-value 0.002 is less than 0.025) in SBP. In terms of DBP, there are statistically significant disparities between men and women in Asmara, Massawa, and Keren but not in Mandafra or Eritrea.

**Table 1:** Socio-demographic and basic background characteristics of the study participants for continuous variables stratified by gender

| Continuous variable | Combined | Male | Female |
| --- | --- | --- | --- |
| **Age (Years)** |  |  |  |
| Mean (SD) | 39.69 (12.38) | 37.00 (13.24) | 41.16 (11.58) |
| Median (IQR) | 40.00 (21) | 34.50 (25) | 42.00 (19) |
| **Height (m)** |  |  |  |
| Mean (SD) | 1.62 (0.09) | 1.70 (0.07) | 1.57 (0.06) |
| Median (IQR) | 1.61 (0.13) | 1.7 (0.10) | 1.57 (0.09) |
| **Weight (kg)** |  |  |  |
| Mean (SD) | 58.85 (12.23) | 61.81 (12.09) | 57.24 (12.01) |
| Median (IQR) | 57.9 (16) | 60.10 (15) | 56 (16) |
| **Body Mass index (kg/m2)** |  |  |  |
| Mean (SD) | 22.49 (4.56) | 21.36 (4.12) | 23.1 (4.65) |
| Median (IQR) | 22.03 (6.2) | 21.02 (5) | 22.72 (6.7) |
| **Sleeping Hours (per day)** |  |  |  |
| Mean (SD) | 7.75 (0.86) | 7.68 (0.81) | 7.78 (0.88) |
| Median (IQR) | 8 (0.0) | 8 (0.0) | 8 (0.0) |

**SD: Standard deviation, IQR: Interquartile range**

**Table 2:** Socio-demographic and basic background characteristics of the study participants for categorical variables stratified by gender

| Categorical Variable | Combined |  | Male |  | Female |  |
| --- | --- | --- | --- | --- | --- | --- |
|  | N | % | N | % | N | % |
| **City** |  |  |  |  |  |  |
| Asmara | 401 | 42.4 | 128 | 38.7 | 273 | 44.7 |
| Massawa | 163 | 17.3 | 74 | 22.4 | 87 | 14.2 |
| Mendefera | 177 | 18.7 | 50 | 15 | 127 | 20.8 |
| Keren | 204 | 21.6 | 79 | 23.9 | 124 | 20.3 |
| **Education** |  |  |  |  |  |  |
| Illiterate | 153 | 16 | 26 | 7.3 | 127 | 20.8 |
| Primary | 137 | 14.5 | 28 | 8.5 | 108 | 17.7 |
| Junior | 252 | 26.7 | 66 | 19.9 | 186 | 30.4 |
| Secondary | 277 | 29.3 | 120 | 36.3 | 157 | 25.7 |
| College | 123 | 13 | 92 | 27.8 | 31 | 5.1 |
| **Marital Status** |  |  |  |  |  |  |
| Married | 659 | 69.7 | 197 | 59.5 | 461 | 75.5 |
| Widowed | 24 | 2.5 | 1 | 0.3 | 23 | 3.8 |
| Divorced | 25 | 2.6 | 1 | 0.3 | 24 | 3.9 |
| Separated | 5 | 0.5 | 0 | 0 | 5 | 0.8 |
| Single/Never married | 226 | 23.9 | 131 | 39.6 | 95 | 15.5 |
| **Ethnic Group** |  |  |  |  |  |  |
| Afar | 27 | 2.9 | 13 | 3.9 | 14 | 2.3 |
| Blen | 87 | 9.2 | 38 | 11.5 | 49 | 8 |
| Nara | 1 | 0.1 | 1 | 0.3 |  |  |
| Saho | 29 | 3.1 | 8 | 2.4 | 20 | 3.3 |
| Tigre | 142 | 15 | 76 | 23 | 66 | 10.8 |
| Tigrigna | 656 | 69.4 | 195 | 58.9 | 461 | 75.5 |
| **Exercise** |  |  |  |  |  |  |
| Yes | 172 | 18.3 | 119 | 36.1 | 53 | 8.7 |
| No | 770 | 81.7 | 211 | 63.9 | 558 | 91.3 |
| **BMI** |  |  |  |  |  |  |
| Underweight | 141 | 14.9 | 59 | 17.8 | 82 | 13.4 |
| Normal weight | 534 | 56.5 | 209 | 63.1 | 324 | 53 |
| Overweight | 207 | 21.9 | 53 | 16 | 154 | 25.2 |
| Obese | 63 | 6.7 | 10 | 3 | 51 | 8.3 |
| **Sleeping Hours (per day)** |  |  |  |  |  |  |
| Less than 6 hours | 25 | 2.6 | 10 | 3 | 15 | 2.5 |
| 6-8 hours | 849 | 89.9 | 301 | 90.9 | 547 | 89.5 |
| More than 8 hours | 62 | 6.6 | 13 | 3.9 | 49 | 8 |

**Table 3:** Mann-Whitney test to examine the study variables difference according to the gender.

| Region | Variable | Gender | N | Median | Mann-Whitney U | P-value |
| --- | --- | --- | --- | --- | --- | --- |
| Eritrea | SBP | Male | 330 | 120 | 98896 | 0.786 |
|  |  | Female | 604 | 118 |  |  |
|  | DBP | Male | 330 | 78 | 94440 | 0.180 |
|  |  | Female | 604 | 78 |  |  |
| Asmara | SBP | Male | 127 | 120 | 15092 | 0.053 |
|  |  | Female | 207 | 118 |  |  |
|  | DBP | Male | 127 | 80 | 14595 | 0.016 |
|  |  | Female | 270 | 76 |  |  |
| Massawa | SBP | Male | 74 | 119 | 2891 | 0.315 |
|  |  | Female | 86 | 114 |  |  |
|  | DBP | Male | 74 | 80 | 2582 | 0.038 |
|  |  | Female | 86 | 74 |  |  |
| Mandafra | SBP | Male | 49 | 120 | 2941 | 0.437 |
|  |  | Female | 127 | 120 |  |  |
|  | DBP | Male | 49 | 80 | 3069 | 0.884 |
|  |  | Female | 127 | 78 |  |  |
| Keren | SBP | Male | 80 | 110 | 3923 | 0.022 |
|  |  | Female | 121 | 118 |  |  |
|  | DBP | Male | 80 | 73 | 3701 | 0.004 |
|  |  | Female | 121 | 80 |  |  |

The findings of the Kruskal-Wallis test, which looked at how the research variables varied by location, are presented in table 4. The findings show that neither the SBP nor the DBP had statistically significant differences between areas (p-values greater than 0.05 in the two cases).

**Table 4:** Kruskal-Wallis test to examine the study variables difference according to the region.

| Variable | Region | N | Median | Chi-Square | P-value |
| --- | --- | --- | --- | --- | --- |
| SBP | Asmara | 397 | 120 | 3.298 | 0.348 |
|  | Massawa | 161 | 117 |  |  |
|  | Mandafra | 176 | 120 |  |  |
|  | Keren | 201 | 117 |  |  |
| DBP | Asmara | 397 | 78 | 0.652 | 0.905 |
|  | Massawa | 161 | 76 |  |  |
|  | Mandafra | 176 | 80 |  |  |
|  | Keren | 201 | 76 |  |  |

In all regions, and especially in Massawa, the scatter plots in figure 3 demonstrate a positive linear correlation between SBP and age. In contrast, age has no bearing on DBP, with data distribution being constant from 20 to 60 years of age.

The statistics in Table 5 detail the SBP and DBP reference levels in Eritrea. The findings show that SBP has a mean and median of 118, a standard deviation of 16.3, and an interquartile range of 17. Between 90 at the first percentile and 170 at the 99th percentile, the percentiles were spread. These findings show that 90% of the population has SBP between 95 (fifth percentile) and 150, while 95% of the population has SBP between 90 (2.5^th^ percentile) and 159 (97.5^th^ percentile) (95th percentile). Moreover, 10% of the population has SBP less than or equal to 100, and 10% has SBP greater than 140 (90^th^ percentile).

**Table 5:** descriptive statistics of SBP and DBP reference values.

| Variable | Measurements | Asmara | Massawa | Mandafra | Keren | Eretria |
| --- | --- | --- | --- | --- | --- | --- |
| SBP | N | 393 | 158 | 175 | 194 | 121 |
|  | Mean (St.D) | 119 (16.6) | 119 (18) | 117 (15) | 116 (15) | 118 (16.3) |
|  | Median (IQR) | 120 (20) | 117 (20) | 120 (12) | 117 (20) | 118 (17) |
|  | P1 | 90 | 88 | 83 | 90 | 90 |
|  | P2.5 | 90 | 90 | 90 | 92 | 90 |
|  | P5 | 98 | 91 | 90 | 95 | 95 |
|  | P10 | 100 | 100 | 100 | 100 | 100 |
|  | P25 | 110 | 110 | 110 | 105 | 110 |
|  | P50 | 120 | 117 | 120 | 117 | 118 |
|  | P75 | 130 | 130 | 122 | 125 | 128 |
|  | P90 | 140 | 140 | 140 | 134 | 140 |
|  | P95 | 148 | 160 | 144 | 142 | 150 |
|  | P97.5 | 154 | 170 | 150 | 157 | 159 |
|  | P99 | 164 | 174 | 169 | 160 | 170 |
| DBP | N | 393 | 158 | 175 | 194 | 121 |
|  | Mean (St.D) | 76 (9.9) | 77 (10.6) | 76 (8.7) | 76 (9.2) | 76 (9.7) |
|  | Median (IQR) | 78 (12) | 76 (14) | 80 (10) | 76 (10) | 78 (10) |
|  | P1 | 60 | 59 | 58 | 58 | 60 |
|  | P2.5 | 60 | 60 | 60 | 60 | 60 |
|  | P5 | 60 | 60 | 60 | 60 | 60 |
|  | P10 | 60 | 62 | 64 | 65 | 62 |
|  | P25 | 70 | 70 | 70 | 70 | 70 |
|  | P50 | 78 | 76 | 80 | 76 | 78 |
|  | P75 | 82 | 84 | 80 | 80 | 80 |
|  | P90 | 88 | 90 | 86 | 88 | 90 |
|  | P95 | 90 | 96 | 90 | 90 | 90 |
|  | P97.5 | 94 | 100 | 92 | 100 | 96 |
|  | P99 | 102 | 108 | 102 | 102 | 102 |

Moreover, DBP had a mean and median of 96 and 78, respectively. 90% of people have DBPs between 60 (fifth percentile) and 90, while 95% of people have DBPs between 60 (2.5th percentile) and 96 (97.5th percentile) (95th percentile). 10% of people have DBP greater than 90, whereas 10% have DBP less than or equal to 62 (10th percentile) (90th percentile).

## Discussion

In our study, we found a significantly less diastolic blood pressure between females in comparison to males in Asmara, Massawa and keren cities, several factors could potentially contribute to this observed difference. Firstly, hormonal variations between males and females, such as the influence of estrogen and testosterone, may play a role in regulating blood pressure. Estrogen, for instance, has been shown to have a vasodilatory effect through increases the gene expression of vasodilatory enzymes such as nitric oxide synthase (NOS) and prostacyclin synthase (13), potentially resulting in lower blood pressure levels in females compared to males. Mishra et al (14) findings indicate that testosterone acts as a facilitating factor in the progression of hypertension triggered by Ang II. This implies that testosterone have a permissive effects on Ang II. Furthermore, the study suggests that testosterone enhances the responsiveness of blood vessels to vasoconstrictor agents, which further exacerbates hypertension.

Our study findings indicate that there are no statistically significant differences in diastolic blood pressure levels among the cities of Asmara, Massawa, Mandafara and Keren. These results suggest that the diastolic blood pressure measurements obtained from participants in these cities do not exhibit substantial variations that can be attributed to the geographical location or urban setting. In our study, we observed that the normal blood pressure values among Eritreans were comparable to those of Sudanese(15), Ethiopians(16), and international standards for blood pressure. These findings indicate that the blood pressure measurements obtained from the Eritrean population align with the expected values observed in neighboring populations and are consistent with internationally recognized standards for blood pressure levels.

## Data Availability

All data produced in the present study are available upon reasonable request to the authors

## Author contributions

Project administration, methodology review & editing were contributed by Ahmed O Noury, O. A. Musa. Data collections were contributed by Ahmed O Noury, Barakat. Data analysis and interpretation were contributed by Ahmed O Noury and Montasir. Manuscript writing was contributed by Ahmed O Noury. Final approval of manuscript was contributed by all authors. All authors read and approved the final manuscript.

## Availability of data and materials

Data supporting this study are included within the article.

## Declarations

### Ethics approval and consent to participate

The Eritrean Ministry of Health’s ethics committee granted ethical approval for this study.

### Consent for publication

All authors have agreed to publish this manuscript.

## References

1. Rehman S, Nelson VL. Blood Pressure Measurement. In: StatPearls [Internet]. StatPearls Publishing; 2021.

2. Flack JM, Adekola B. Blood pressure and the new ACC/AHA hypertension guidelines. Trends Cardiovasc Med. 2020;30(3):160–4.

3. Whelton PK, Carey RM, Aronow WS, Casey DE, Collins KJ, Dennison Himmelfarb C, et al. 2017 ACC/AHA/AAPA/ABC/ACPM/AGS/APhA/ASH/ASPC/NMA/PCNA guideline for the prevention, detection, evaluation, and management of high blood pressure in adults: a report of the American College of Cardiology/American Heart Association Task Force on Clinical Practice Guidelines. J Am Coll Cardiol. 2018;71(19):e127–248.

4. Williams B, Mancia G, Spiering W, Agabiti Rosei E, Azizi M, Burnier M, et al. 2018 ESC/ESH Guidelines for the management of arterial hypertension: The Task Force for the management of arterial hypertension of the European Society of Cardiology (ESC) and the European Society of Hypertension (ESH). Eur Heart J. 2018;39(33):3021–104.

5. Lindhorst J, Alexander N, Blignaut J, Rayner B. Differences in hypertension between blacks and whites: an overview. Cardiovasc J Afr. 2007;18(4):241–7.

6. Te Riet L, van Esch JHM, Roks AJM, van den Meiracker AH, Danser AHJ. Hypertension: renin–angiotensin–aldosterone system alterations. Circ Res. 2015;116(6):960–75.

7. Feelisch M, Kolb-Bachofen V, Liu D, Lundberg JO, Revelo LP, Suschek C V, et al. Is sunlight good for our heart? Eur Heart J. 2010;31(9):1041–5.

8. Opländer C, Deck A, Volkmar CM, Kirsch M, Liebmann J, Born M, et al. Mechanism and biological relevance of blue-light (420–453 nm)-induced nonenzymatic nitric oxide generation from photolabile nitric oxide derivates in human skin in vitro and in vivo. Free Radic Biol Med. 2013;65:1363–77.

9. Liu D, Fernandez BO, Hamilton A, Lang NN, Gallagher JMC, Newby DE, et al. UVA irradiation of human skin vasodilates arterial vasculature and lowers blood pressure independently of nitric oxide synthase. J Invest Dermatol. 2014;134(7):1839–46.

10. Alvo M. Multivariate Methods. In: Statistical Inference and Machine Learning for Big Data. Springer; 2022. p. 63–93.

11. Kabacoff R. R in Action: Data Analysis and Graphics with R and Tidyverse. Simon and Schuster; 2022.

12. Kvam P, Vidakovic B, Kim S. Nonparametric Statistics with Applications to Science and Engineering with R. Vol. 1. John Wiley & Sons; 2022.

13. Siow RCM, Li FYL, Rowlands DJ, de Winter P, Mann GE. Cardiovascular targets for estrogens and phytoestrogens: transcriptional regulation of nitric oxide synthase and antioxidant defense genes. Free Radic Biol Med. 2007;42(7):909–25.

14. Mishra JS, More AS, Gopalakrishnan K, Kumar S. Testosterone plays a permissive role in angiotensin II-induced hypertension and cardiac hypertrophy in male rats. Biol Reprod. 2019;100(1):139–48.

15. Musa OA, Elnagi Y. Hago. Normal Blood Pressure in Adult Sudanese in Khartoum State, Sudan.

16. Beall CM, Gebremedhin A, Brittenham GM, Decker MJ, Shamebo M. Blood pressure variation among Ethiopians on the Simien Plateau. Ann Hum Biol. 1997;24(4):333–42.

